# Underlying factors that influence the acceptance of COVID-19 vaccine in a country with a high vaccination rate

**DOI:** 10.1101/2021.10.31.21265676

**Authors:** Daniela Toro-Ascuy, Nicolás Cifuentes-Muñoz, Andrea Avaria, Camila Pereira-Montecinos, Gilena Cruzat, Francisco Zorondo-Rodriguez, Loreto F Fuenzalida

## Abstract

Control of the COVID-19 pandemic largely depends on the effectiveness of the vaccination. Several factors including vaccine hesitancy can affect the vaccination process. Understanding the factors that underlie the willingness to accept vaccination brings pivotal information to control the pandemic. We analyzed the association between the willingness level to accept the COVID-19 vaccine, and vaccine determinants amidst the Chilean vaccination process. Individual-level survey data was collected from nationally representative samples of 744 respondents, and multivariate regression models used to estimate the association between outcome and explanatory variables. We found that trust in the COVID-19 vaccine, scientists, and medical professionals increased the willingness to: accept the vaccine, a booster dose, annual vaccination, and children vaccination. Our results are critical to understand the acceptance of COVID-19 vaccines in the context of a country with one of the world’s highest vaccination rates. We provide information for decision-making, policy design and communication of vaccination programs.

## Introduction

The sudden entry of the severe acute respiratory syndrome coronavirus 2 (SARS-CoV-2) into the human population in 2019 has had catastrophic consequences, with global deaths over 4.8 million worldwide^1,2^. In March 2020, the World Health Organization declared the coronavirus disease 2019 (COVID-19) a pandemic. In Chile, the first reported case of COVID-19 occurred in March 2020. Community transmission of the virus caused the peak of the first wave of COVID-19 cases in June 2020^3^. Non-pharmaceutical interventions were key to slow down the pandemic in Chile and prevent further deaths, which account for over 37,000 as of October 2021.

Worldwide, non-pharmaceutical interventions such as mask use, lockdowns and social distancing helped to slow down the pandemic until vaccines became available. The remarkably rapid development of vaccines against SARS-CoV-2 is turning COVID-19 into a preventable disease^4^. However, several challenges about COVID-19 vaccination remain to be addressed, including the hesitance to accept the vaccines. The first vaccine approved for emergency use in Chile was the Pfizer-Biontech BNT162-b2 on December 16, 2020; The second vaccine to be approved by the Chilean authorities for emergency use was the virion-inactivated Sinovac vaccine, on January 20 of 2021. Sinovac has been the most widely used vaccine in Chile, with over 20 million doses administered, followed by BNT162-b2 with over 7 million doses, by the end of September 2021. Studies have shown a high effectiveness of Sinovac in preventing symptomatic COVID-19, hospitalizations and death^5^. Recently, emergency use authorization has been granted for the AZD1222 (Oxford-AstraZeneca), Ad26.COV2.S/Janssen (Johnson & Johnson), Ad5-nCoV/CanSino and GAM-COVIDVAC (Gamaleya) in Chile.

The success of vaccination strongly depends on underlying social factors, mainly willing to accept the vaccination, trust in stakeholders related to vaccination, vaccine-specific factors, communication and media, historical influences, religion, gender, socioeconomic, politics, geographic barriers, experience with vaccination, risk perception, and design of the vaccination program^6^. Some studies explored COVID-19 vaccine acceptance and their determinants using surveys in different countries including the United States^7-9^, United Kingdom^9-11^, China^12,13^, Indonesia^14^, Italy^15^, Ireland^10^and Japan^16^. In addition, surveys have explored vaccine acceptance in groups of European countries^17^, Arab countries^18,19^ and other countries worldwide^20^. Several of these studies have concluded that the willingness to accept the vaccine differs depending on the age, educational and economic level, credibility in government decisions and perception of risk of COVID-19 disease^7,12,18,20-22^. In a global vaccine study carried out in 19 countries, it was reported that responses have had high heterogeneity depending on the country surveyed^20^; therefore, it is important to understand the acceptance of a vaccine in each country or region^22^. Vaccination against SARS-CoV-2 remains a big challenge for most of the countries, especially those with poor economies. Countries with broad vaccination coverage can offer key lessons on how to address the challenges about COVID-19 vaccination. In this context, the Chilean COVID-19 vaccination campaign has emerged as one of the most successful and rapid worldwide^23^. By July 2021, Chile was among the first countries in the world with more doses administered per 100 people (our world in data). However, as observed in Chile and many other countries, vaccination has slowed down after reaching over 70% of the population fully immunized.

Understanding the determinants of vaccine acceptance is key for decision-making, for establishing differentiated strategies according to the characteristics and social determinations of the population, and for identifying the subjectivities that underlie the decision-making of the people to vaccinate. Faced with the need to obtain scientific evidence amidst the Chilean vaccination campaign, we analyzed the association between the level of willingness to accept the COVID-19 vaccine and determinants of the vaccine in Chile. We focused on trust in vaccines and stakeholders and perceptions of people about effectiveness of prevention practices, infection risk, and side effects, to estimate their associations with willingness to accept the SARS-CoV-2 vaccine, booster dose, annual vaccination, and vaccination of children. Our study identified several key aspects such as a high trust in scientists and health area workers, as well as a moderate trust in the media. Our results permit projecting subjective dimensions related to the decision of the population to be vaccinated, identifying the risks and trusts of the Chilean population associated with this process, which offer keystone evidence for other countries to face the pandemic. We identified several aspects such as perception of risk and prevention practice, and trust related with the vaccination process, scientists and medical professionals, and sociodemographic variables associated with the acceptance of vaccination, booster doses, or children vaccination. The information provided by our study is relevant to improve public health communication strategies.

## Results

Between May 21 and June 21 of 2021, a total of 744 adults were recruited in Chile via online surveys. The self-application questionnaire was disseminated through social networks. A summary of the socio-demographic characteristics of respondents included in this study is shown in Table S1. The questions were aimed at estimating four outcome variables related to willing to accept: (i) SARS-CoV-2 vaccination (0=not, 1=maybe, 2=yes), (ii) vaccine booster dose (1=yes), (iii) annual vaccination (1=totally disagree to 4=totally agree), and (iv) vaccination of children (1=totally disagree to 4=totally agree). The questionnaire also captured a set of social determinants and attributes associated with the vaccine as explanatory variables. The questions for explanatory variables were aimed at describing the perception of risk, trust, and their responses also were recorded at a level of agreement or disagreement, ordered on a 3 or 4-point ordinal scale. For instance, trust in the different stakeholders about COVID-19 vaccines was estimated using a scale: “No trust”, “Little trust”, “Pretty trust”, “High trust”. The full questionnaire is shown in the Supplementary Information (Table S2). In addition, Table S2 indicates milestones that occurred during the data collection period, according to the development of the pandemic and vaccination in Chile. Most of the respondents (93.4% n= 695) had received at least one dose of SARS-CoV-2 vaccine at the time of the survey, whereas 3.9% (n=29) still had not decided if they would accept a SARS-CoV-2 vaccine and 2.7% (n=20) of respondents said they would definitely not accept a SARS-CoV-2 vaccine. 88.2% (n=656) of the respondents reported that they would accept an hypothetical booster dose and 57.8% (n=430) said they would definitively accept to get vaccinated every year if necessary, similar to the vaccine schedule of influenza virus. When asked if in case of having children under 16 years old, they would accept that their children could be vaccinated against SARS-CoV-2, 62.5% (n=175) reported that they would “definitively accept” a SARS-CoV-2 vaccine for their children.

Multivariate regression models were used to estimate the association between outcome and explanatory variables. Ordered logistic regression model was adjusted when analyzing the outcome variables of willingness to accept the SARS-CoV-2 vaccination, annual vaccination, and vaccination of children, while logistic regression model was used to analyze the willingness to accept the vaccine booster dose. For each outcome variable, we selected the model with best goodness of fit and parsimony using Akaike information criterion (AIC) (Table S3).

We computed the odds ratio for selected models, which represents the ratio of the odds that an outcome variable will occur given an explanatory variable compared to the odds of the outcome occurring in the absence of the explanatory variable. If the odd ratio is greater than 1, then the explanatory variable induces a higher level of acceptance, relative to the control of other variables used in the model. Rather, an odd ratio smaller than 1 suggests that an explanatory variable influences a lower willingness. We described only those results where the 95% confidence intervals exclude zero, which were deemed statistically credible. Although our models do not measure causal effects, log cumulative odds ratios show how the variables on willingness respond to variables of perception and trust or how the associations would vary between genders or age groups. The access to self-reported perceptions provide correlational evidence of which underlying factors could explain a greater willingness.

### Trust in SARS-CoV-2 vaccine increased the willingness to accept the SARS-CoV-2 vaccine, booster dose, annual vaccination and vaccination of children

The trust of people varies between vaccines (Kruskal-Wallis test: Chi-squared=509, d.f.=5, p<0.001). While reported trust did not differ between Sinovac and Pfizer vaccines (Dunn’s test with Bonferroni adjustment: z=-1.77, p=0.57), the trust in Sinovac and Pfizer vaccines were significantly greater than trusts in the other four vaccines approved by Chilean authorities (Dunn’s test with Bonferroni adjustment between Sinovac vs. CanSino z=12.5, p<0.001; vs. AstraZeneca z=12.9, p<0.001; vs. Sputnik z=14.5, p<0.001; vs. Johnson z=13.0, p<0.001; and between Pfizer vs. CanSino z=14.1, p<0.001; vs. AstraZeneca z=14.6, p<0.001; vs. Sputnik z=16.1, p<0.001; vs. Johnson z=14.6, p<0.001). There were no differences in the reported trust in the vaccines of CanSino, AstraZeneca, Sputnik and Johnson. In spite of differences between trust in vaccines, trust of all vaccines is retained in one factor (Factor Analysis: Eigenvalue of Factor 1=3.6; LR test: chi-squared=2032.1, p<0.001) with a very high reliability coefficient (Cronbach’s alpha=0.89). So, we took the average of trust of all vaccines to create a variable of overall trust of vaccines against SARS-CoV-2 and used it in the multivariate regression models.

Multivariate models suggest that the increase of one unit value in trust of SARS-CoV-2 vaccines increased 4.1 times the willing to accept the SARS-CoV-2 vaccines (95%CI=2.0-8.2, p<0.001), 3.2 times the probability of the willing to accept a booster dose (95%CI=1.8-5.6, p<0.001), twice the willing to accept annual vaccination (95%CI=1.6-2.8, p<0.001), and 1.9 times the willing to vaccinate children (95%CI=1.4-2.6, p<0.001) (Table 1, row [a]). When comparing results between genders, women showed significant associations of all willingness variables with trust in SARS-CoV-2 vaccine (Table S4). On the other hand, men showed a significant association between willingness to accept the annual vaccination and trust in SARS-CoV-2 vaccine (Table S4). Interestingly, it was observed in both young people and adults that an increase of trust in SARS-CoV-2 vaccine would induce a higher willingness toward SARS-CoV-2 vaccination, booster dose, annual vaccination and vaccination of children (Table S5).

**Table 1.**
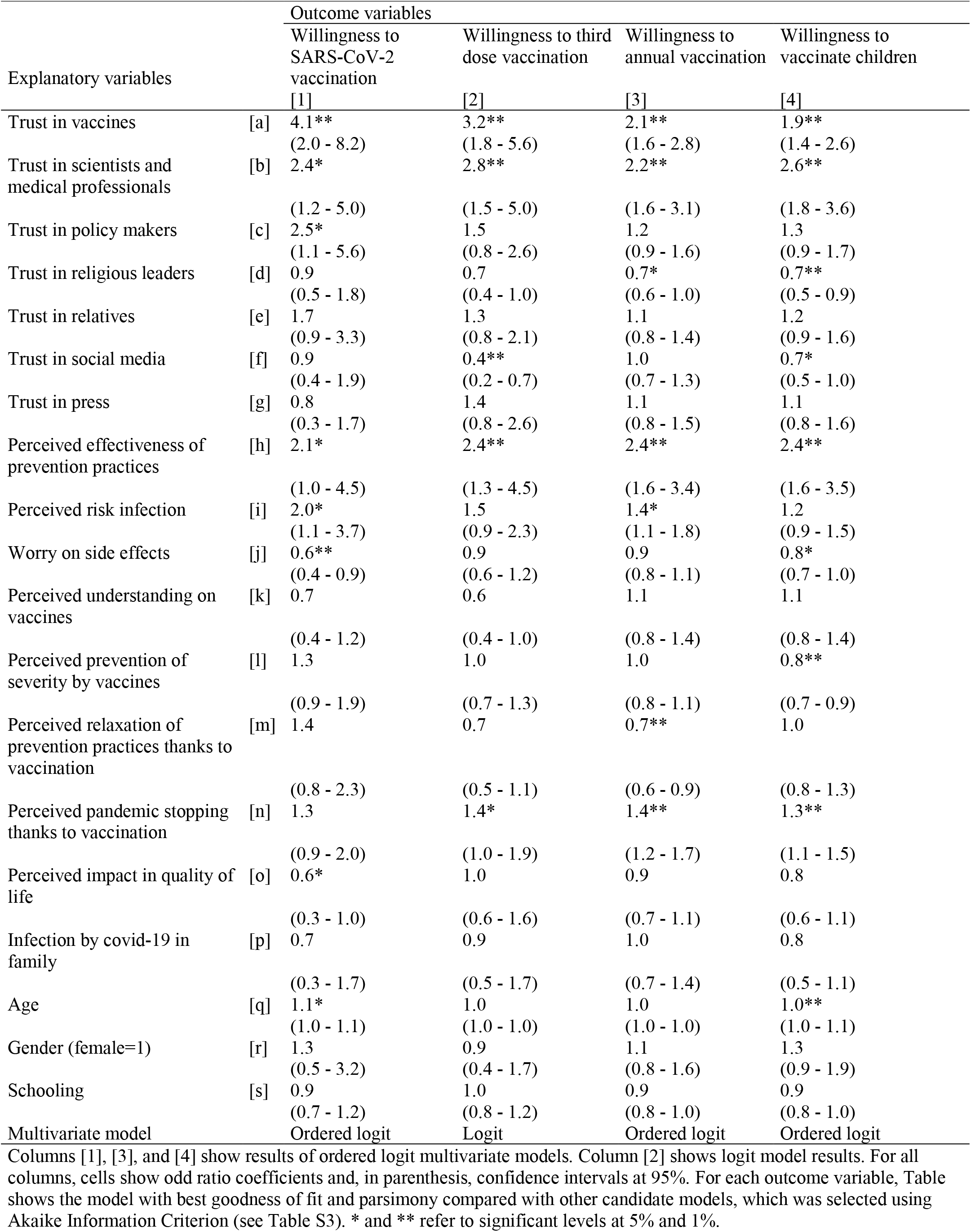
Associations of the willingness to SARS-CoV-2 vaccination, third dose, annual vaccination, and to vaccinate children, with variables of trust and perception among Chileans (n=744, 2021).

| Explanatory variables |  | Outcome variables |  |  |  |
| --- | --- | --- | --- | --- | --- |
|  |  | Willingness to SARS-CoV-2 vaccination | Willingness to third dose vaccination | Willingness to annual vaccination | Willingness to vaccinate children |
|  |  | [1] | [2] | [3] | [4] |
| Trust in vaccines | [a] | 4.1**<br>(2.0 - 8.2) | 3.2**<br>(1.8 - 5.6) | 2.1**<br>(1.6 - 2.8) | 1.9**<br>(1.4 - 2.6) |
| Trust in scientists and medical professionals | [b] | 2.4*<br>(1.2 - 5.0) | 2.8**<br>(1.5 - 5.0) | 2.2**<br>(1.6 - 3.1) | 2.6**<br>(1.8 - 3.6) |
| Trust in policy makers | [c] | 2.5*<br>(1.1 - 5.6) | 1.5<br>(0.8 - 2.6) | 1.2<br>(0.9 - 1.6) | 1.3<br>(0.9 - 1.7) |
| Trust in religious leaders | [d] | 0.9<br>(0.5 - 1.8) | 0.7<br>(0.4 - 1.0) | 0.7*<br>(0.6 - 1.0) | 0.7**<br>(0.5 - 0.9) |
| Trust in relatives | [e] | 1.7<br>(0.9 - 3.3) | 1.3<br>(0.8 - 2.1) | 1.1<br>(0.8 - 1.4) | 1.2<br>(0.9 - 1.6) |
| Trust in social media | [f] | 0.9<br>(0.4 - 1.9) | 0.4**<br>(0.2 - 0.7) | 1.0<br>(0.7 - 1.3) | 0.7*<br>(0.5 - 1.0) |
| Trust in press | [g] | 0.8<br>(0.3 - 1.7) | 1.4<br>(0.8 - 2.6) | 1.1<br>(0.8 - 1.5) | 1.1<br>(0.8 - 1.6) |
| Perceived effectiveness of prevention practices | [h] | 2.1*<br>(1.0 - 4.5) | 2.4**<br>(1.3 - 4.5) | 2.4**<br>(1.6 - 3.4) | 2.4**<br>(1.6 - 3.5) |
| Perceived risk infection | [i] | 2.0*<br>(1.1 - 3.7) | 1.5<br>(0.9 - 2.3) | 1.4*<br>(1.1 - 1.8) | 1.2<br>(0.9 - 1.5) |
| Worry on side effects | [j] | 0.6**<br>(0.4 - 0.9) | 0.9<br>(0.6 - 1.2) | 0.9<br>(0.8 - 1.1) | 0.8*<br>(0.7 - 1.0) |
| Perceived understanding on vaccines | [k] | 0.7<br>(0.4 - 1.2) | 0.6<br>(0.4 - 1.0) | 1.1<br>(0.8 - 1.4) | 1.1<br>(0.8 - 1.4) |
| Perceived prevention of severity by vaccines | [l] | 1.3<br>(0.9 - 1.9) | 1.0<br>(0.7 - 1.3) | 1.0<br>(0.8 - 1.1) | 0.8**<br>(0.7 - 0.9) |
| Perceived relaxation of prevention practices thanks to vaccination | [m] | 1.4<br>(0.8 - 2.3) | 0.7<br>(0.5 - 1.1) | 0.7**<br>(0.6 - 0.9) | 1.0<br>(0.8 - 1.3) |
| Perceived pandemic stopping thanks to vaccination | [n] | 1.3<br>(0.9 - 2.0) | 1.4*<br>(1.0 - 1.9) | 1.4**<br>(1.2 - 1.7) | 1.3**<br>(1.1 - 1.5) |
| Perceived impact in quality of life | [o] | 0.6*<br>(0.3 - 1.0) | 1.0<br>(0.6 - 1.6) | 0.9<br>(0.7 - 1.1) | 0.8<br>(0.6 - 1.1) |
| Infection by covid-19 in family | [p] | 0.7<br>(0.3 - 1.7) | 0.9<br>(0.5 - 1.7) | 1.0<br>(0.7 - 1.4) | 0.8<br>(0.5 - 1.1) |
| Age | [q] | 1.1*<br>(1.0 - 1.1) | 1.0<br>(1.0 - 1.0) | 1.0<br>(1.0 - 1.0) | 1.0**<br>(1.0 - 1.1) |
| Gender (female=1) | [r] | 1.3<br>(0.5 - 3.2) | 0.9<br>(0.4 - 1.7) | 1.1<br>(0.8 - 1.6) | 1.3<br>(0.9 - 1.9) |
| Schooling | [s] | 0.9<br>(0.7 - 1.2) | 1.0<br>(0.8 - 1.2) | 0.9<br>(0.8 - 1.0) | 0.9<br>(0.8 - 1.0) |
| Multivariate model |  | Ordered logit | Logit | Ordered logit | Ordered logit |

### Trust in scientists and medical professionals increases willingness of SARS-CoV-2 vaccination, booster dose, annual vaccination, and children vaccination, while trust in religious leaders would reduce the willingness to accept the annual vaccination and vaccination of children

The people’s trust varied significantly between stakeholders (Kruskal-Wallis test: 2285.9, d.f.=8, p<0.001). Scientists received the highest score of trust among all stakeholders included in the study (Dunn’s test with Bonferroni adjustment: z>6.7 and p<0.001 in all comparisons), followed by medical professionals, Chilean Public Health Institute (ISP), and WHO’s professionals. The lowest scores of trusts were reported for policy makers and religious leaders. For instance, 63% (n=472) and 28% (n=207) of individuals reported to have a “high” or “pretty trust”, respectively, in scientists, and more than 70% of people reported to have a “high” trust (43%, n=323) or “pretty trust” (36%, n=267) in medical professionals. On the contrary, 64% (n=479) and 45% (n=334) of the individuals reported to not trust in religious leaders and policy makers. Factor analysis suggests that the variability of trusts in stakeholders included in the study can be explained by four groups: (a) scientists and medical professionals group, that includes trust in scientists and medical, WHO’s, and ISP’s professionals (Retained Factor 1: Eigenvalue=2.3, LR test Chi-square=1407, p<0.001, Cronbach’s alpha=0.85); (b) Policy makers group that includes policy makers and authorities of the Ministry of Health (Retained Factor 1, Eigenvalue=0.8; LR test Chi-square=247.3, p<0.001; Cronbach’s alpha=0.7); (c) Relatives group includes relatives and friends (Retained Factor 1=0.8, LR test Chi-Square=289.9, p<0.001; Cronbach’s alpha=0.7), and (d) a fourth group with religious leaders only. To include trust in stakeholder groups in the multivariate regression models, we averaged the reported trust scores for all stakeholders included in each group.

We found evidence that individuals with higher trust in scientists and medical professionals significantly increased 2.4 times their willingness toward SARS-CoV-2 vaccination (95% CI=1.2-5.0, p=0.01) as well as 2.8 times their willingness to accept the booster dose (95% CI=1.5-5.0, p=0.001) (Table 1, row [b] of the columns [1]-[2]). Similarly, an increase of trust in scientists and medical professionals also increased 2.2-fold the willingness to accept annual vaccination (95% CI=1.6-3.1, p<0.001) and 2.6 times the vaccination of children (95% CI=1.8-3.6, p<0.001) (Table 1, row [b] of the columns [3]-[4]). Interestingly, some groups responded differently to trust in scientists and medical professionals and showed comparatively different associations with the willingness variables. For instance, women did not vary their willingness to accept SARS-CoV-2 vaccine and booster dose, when they reported a higher or lower level of trust in scientists and medical professionals. In contrast, men showed that a higher trust in scientists and medical professionals increased 46.1 and 4.2 times the willingness to accept the SARS-CoV-2 vaccine (95% CI=2.5-862.1, p=0.01) and booster dose (95% CI=1.3-13.1, p=0.02), respectively (Table S4). In the case of the willingness to accept the annual vaccination and vaccination of children, both women and men showed a similar positive impact of trust in scientists and medical professionals (Table S4). We found evidence that young and adults differed on how their trust in scientists and medical professionals would impact the variables of willingness. For instance, young people did not vary their willingness to accept the SARS-CoV-2 vaccine, booster dose, annual vaccination, and vaccination of children as their levels of trust in scientists and medical professionals increased (Table S5). Unlike, adults showed that a higher level of trust increases around three-fold the willingness of SAR-CoV-2 vaccine, booster dose, annual vaccination, and vaccination of children (Table S5). In contrast, the results show that willingness to accept both annual vaccination (95%CI=0.6-0.9, p=0.02) and children vaccination (95%CI=0.6-0.9, p=0.004) decreased in 30% as increases one unit of trust in religious leaders (Table 1, row [d] of the columns [3]-[4]). However, when comparing between genders, the trust in religious leaders decreases the willingness to accept the booster dose only among men. Women and men decreased the willingness to vaccinate children as their trust in religious leaders increased (Table S4). Trust in religious leaders differently impacts the willingness scores between age groups. For instance, only adults showed a decrease of the willingness to accept booster dose as trust in religious leaders increases, while only young individuals who have more trust in religious leaders decreased their willingness to accept annual vaccination and vaccination of children.

Last, we also found that an increase of trust in social media is associated with lower willingness to accept booster dose vaccination (OR=0.4, 95%CI=0.2-0.7, p=0.001) and vaccination of children (OR=0.7, 95%CI=0.7-1.0, p=0.03) (Table 1, row [f] of the columns [2] and [4]).

### A higher perceived effectiveness of prevention practices, higher perceived infection risk and being less worried of side effects of vaccines, increased the willingness to accept SAR-CoV-2 vaccine, whilst they are differently associated with the willingness to accept the booster dose, annual vaccination and vaccination of children

People’s perceptions of effectiveness varied across prevention practices (Kruskal-Wallis test: Chi-squared=591.1, d.f.=7, p<0.001). For instance, the vaccination was perceived as more effective compared with lockdown (Dunn’s test with Bonferroni adjustment: z=10.2, p<0.001), but less effective than the use of mask (z=-6.22, p<0.001), washing hands (z=-10.1, p<0.001), physical distance (z=-9.3, p<0.001), avoid meeting people (z=-7.1, p<0.001), and quarantine (z=-3.9, p=0.001). Also, the use of a mask was perceived as less effective than washing hands (z=-4.8, p<0.001) and physical distance (z=-3.3, p=0.011). The lockdown was perceived as the lowest effective practice to prevent the infection of SARS-CoV-2 among all included in the study. Notwithstanding the different perceptions of effectiveness across practices, results suggest that the variability of perceptions can be retained in one factor (Factor Analysis: Eigenvalue of factor 1=3.0; LR test: chi-squared=1292.1, p<0.001) and the set of variables has a high internal consistency (Cronbach’s alpha=0.81). We calculated the overall perceived effectiveness of the prevention practices for each individual as the average value of perceived effectiveness among all practices, and used this new variable in the multivariate analysis.

Most people perceived that the probability of infection with COVID-19 is “little likely” (n=423, 56.8%) or “likely” (n=239, 32.1%) (Fig. 1). Only 49 individuals (6.6%) of the sample reported that getting COVID-19 is “very likely”. In turn, people reported to be “little worried” (n=306, 41%) or “not worried” (n=190, 25.5%) on the side effects of vaccines. A total of 79 individuals of the sample reported to be “very worried” about the side effects of vaccines (n=79, 10.6%) (Fig. 1).

**Figure 1.**
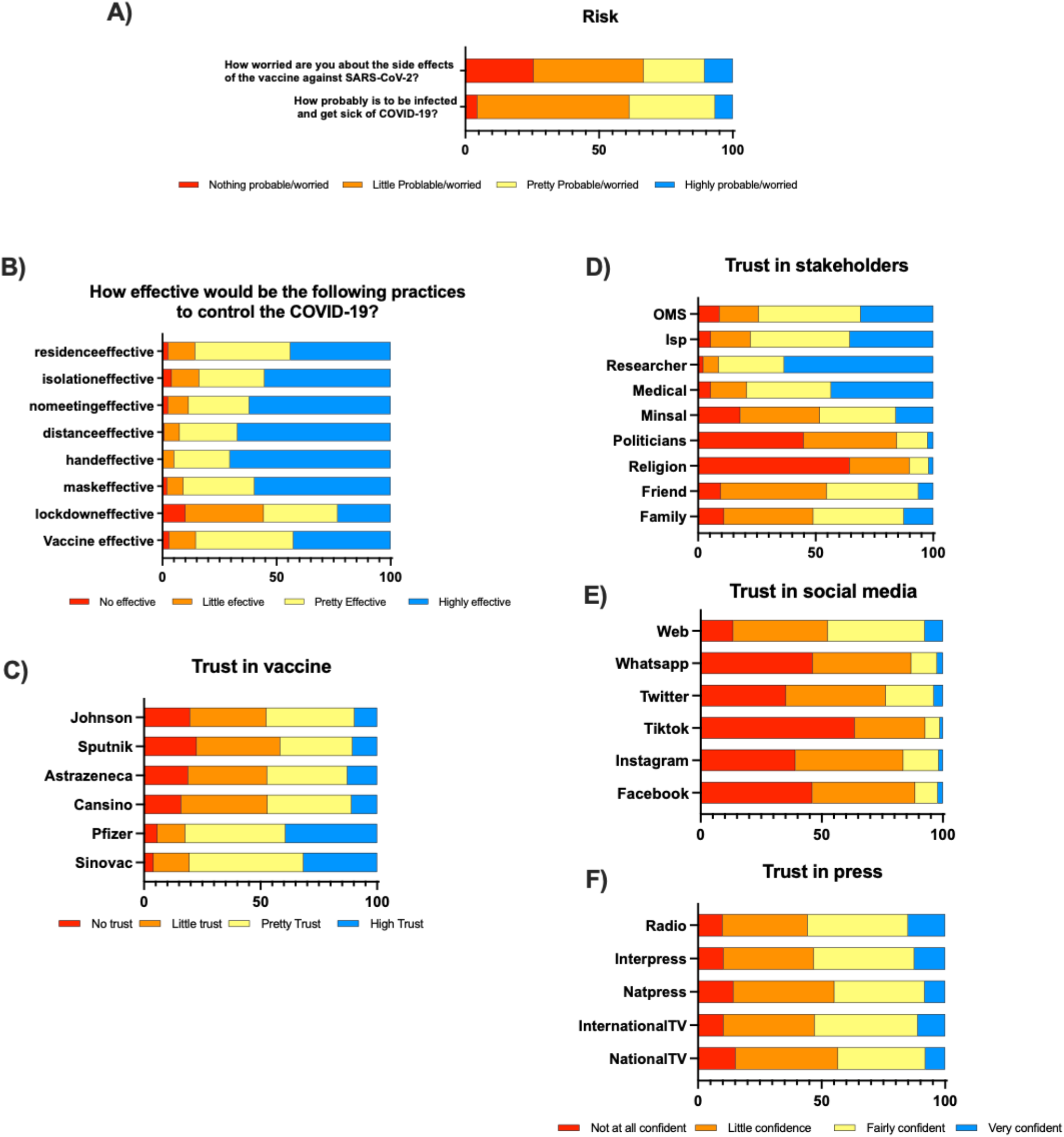
Perceptions of respondents of vaccine side effects, getting sick from COVID-19, effectiveness of COVID-19 control practices, and trust in vaccines, stakeholders, social media, and the press. The bars indicate the breakdown of the percentage of respondents providing an answer given to each question asked. The questionnaire is shown in the Supplementary Information.

A score unit higher in the perceived effectiveness of the practices to prevent infection increased the willingness to accept the SARS-CoV-2 vaccine in 2 fold (95%CI=1.0-4.5, p=0.05), the booster dose in 2.4 times (95%CI=1.3-4.5, p=0.005), annual vaccination in 2.4 times (95%CI=1.6-3.4, p<0.001), and vaccination of children in 2.4 fold (95%CI=1.6-3.5, p<0.001) (Table 1, row [h]). Moreover, the increase in a value unit of perception of infection risk increased twice the willingness to accept SARS-CoV-2 vaccine (95%CI= 1.1-3.7; p=0.02) and 1.4 times the willingness to accept annual vaccination against SARS-CoV-2 (95%CI=1.1-1.8; p=0.01) (Table 1, row [i] of the columns [1] and [3]). In contrast, the increase in a unit value of worry about side effects decreased 40% the willingness to accept SARS-CoV-2 vaccine (95% CI=0.4-0.9; p=0.01) and 20% the willingness to vaccinate children (95% CI=0.7-1.0; p=0.02) (Table 1, row [j] of the columns [1] and [4]).

Moreover, we found that associations of willingness variables with perceived effectiveness, perceived infection risk and to be worried about the side effects, varied between genders and cohorts. For instance, when comparing results between genders, women showed significant associations of all willingness variables with perceived effectiveness of prevention practices (Table S4). On the other hand, men showed a significant association between willingness to accept the annual vaccination and perceived effectiveness of prevention practices (Table S4). Analyses across age groups suggest that only adults increased their willingness to accept the booster dose vaccination as their perceived effectiveness of prevention practices increased. Both adults and young people had shown that an increase in perceived effectiveness increased the willingness to accept the annual vaccination and vaccination of children (Table S5). Also, perception of infection risk was positively and significantly associated with willingness to accept SARS-CoV-2 vaccine (OR=2.4; 95%CI=1.1-5.1; p=0.02) and booster dose (OR=1.7; 95%CI=0.9-3.1; p=0.08) among women, but not among men (Table S4). To be worry about the side effects decreased the willingness to accept SARS-CoV-2 vaccine both among women (OR=0.5; 95% CI=0.3-0.8, p=0.008) and among men (OR=0.2; 95%CI=0.0-0.9; p=0.03), but only men decreased their willingness to vaccinate children as increase their worry about side effects (OR=0.7; 95%CI=0.5-0.9; p=0.02) (Table S4). Similarly, a higher worry about side effects decreased the willingness to accept the SARS-CoV-2 vaccine (OR=0.3, 95%CI=0.1-0.6, p=0.002) and booster dose (OR=0.3, 95%CI=0.1-0.8, p=0.01) only among young people. Instead, adults showed that a higher worry of side effects decreased the willingness to vaccinate children (OR=0.7, 95%CI =0.6-0.9, p=0.01) (Table S5).

Interestingly, when individuals perceived that vaccines reduce the severity of COVID-19, they also increased by 20% their willingness to vaccinate their children (OR=0.8, 95%CI=0.7-0.9, p=0.002) (Table 1, row [l] of the column [4]). A higher perceived relaxation of prevention practices thanks to vaccination was associated with a reduction of the willingness to accept the annual vaccination (OR=0.7, 95%CI=0.6-0.9, p=0.001) (Table 1, row [m] of the columns [3]). Individuals who report a higher agreement that vaccination would stop the pandemic show higher willingness to accept the booster dose (OR=1.4, 95%CI=1.0-1.9, p=0.03), annual vaccination (OR=1.4, 95%CI=1.2-1.7, p<0.001), and vaccination of children (OR=1.3, 95%CI=1.1-1.5, p=0.001) (Table 1, row [n] of the columns [2]-[4]). Last, individuals who report a positive impact of the pandemic in their subjective well-being show a lower willingness to accept the SAR-CoV-2 vaccine (OR=0.6, 95%CI=0.3-1.0, p=0.04), but did not vary their willingness to accept a booster dose, annual vaccination, and vaccination of children (Table 1, row [o]).

## Discussion

The promotion of vaccination against SARS-CoV-2 is key to tackle the COVID-19 pandemic. Vaccination success strongly depends on understanding whether people are willing to accept COVID-19 vaccines, and underlying factors of their acceptance (or refusal). We contribute to the existing literature focusing on whether the willingness to accept the vaccination is impacted by trust and perceptions of different issues related to the COVID-19 pandemic, in the context of a country with one of the world’s highest vaccination rates. Our findings bring relevant information for decision making processes, particularly for the design and communication of vaccination implementation programs in countries where COVID-19 vaccination remains low.

Our results indicate that higher acceptance of the vaccine correlates with a high level of trust in experts of the field (scientists and health workers). Our finding highlights that trust in experts of the field increases the acceptance of SARS-CoV-2 vaccines, booster dose, annual vaccination and vaccination of children. The significant and positive associations emphasize the pivotal role that the trust in experts has on vaccination against COVID-19. In contrast, trust in political or religious leaders is extremely low and on the contrary, when it was high, the refusal to vaccinate was also higher. Our results are consistent with other studies where a high trust in health workers is associated with high acceptance of vaccination^24,25^. This is also consistent with the lack of trust in vaccine experts who have been associated with support towards political attitudes against vaccines^26^. Our findings also underlined that the trusted information and sources are crucial, suggesting it is not only trust but also confidence in health authorities that affects vaccine acceptance^27^.

The trust in the government has been associated with vaccine acceptance^20^. In the Chilean context, trust in national and health authorities is lower^28^ than in other countries^29^. Trust in the Chilean government could have seen diminished within the paradox between the increase in infections despite the success of the vaccination campaign (first doses), likely related to the initial messages of the vaccination campaign from the authorities that placed the vaccine as the only possibility to control the virus^28^. In the Chilean case, all the communes of the capital of Chile entered an obligate quarantine during the data collection period due to the increase in COVID-19 cases. The low trust in the Chilean authorities and its negative impact on willingness to accept vaccination might be the result of generalized criticisms to the authorities due to the management of social inequalities and how the COVID-19 pandemic impacted differently the socioeconomic groups^30^. Our results show the need to deepen this aspect in future studies, as well as to consider the emergence of “new actors’’ within the society in which trust could be deposited, as our findings show: in the scientific world and in the medical personnel itself.

Our study identified that the older population has a higher acceptance in the vaccination process. Higher acceptance of the vaccine in older populations could be associated with the perception of the risk of contagion of SARS-CoV-2. It has been described that the willingness to be vaccinated is more related to age than to gender, indicating that those who perceive a greater susceptibility to the effects of the virus are more open to accept vaccination and even willing to take a booster dose^29^.

The perception of the risk of the pandemic was initially associated with the older age of those affected, and may be related to the perception of the risk of contagion of the virus, as well as the possibility of protecting their health through vaccination.

According to our results, and in concordance with previous studies, trust in vaccines contributes to explain the acceptance of vaccination uptake^31^. In the current SARS-CoV-2 pandemic, the development of new vaccination platforms has been achieved. People reported a high trust in all vaccines, albeit the Sinovac and Pfizer vaccines were the most trusted among people. While Sinovac has reported lower efficacies and effectiveness than Moderna and AstraZeneca vaccines^5,32^ it has been more trusted among Chilean people. High trust in Pfizer and Sinovac vaccines coincides with both of them being the most used in the country. Governments must proactively provide information on selected vaccine manufacturers and new platforms to break down potential barriers of knowledge that may affect trust in vaccines. Consistent with other studies^14,24^, our results strengthen the idea that high trust in vaccines and high perceived effectiveness of prevention practices increase the willingness of people to accept the vaccination for them and their children, as well as to accept the booster dose and annual vaccination programs.

Along this line, our findings suggest that in cases where trust in vaccines and the perception of effectiveness were lower, the authorities must have a leading role in developing strategies to convince, to delve into aspects that relate to what should be communicated, and how to increase confidence and education about the vaccine, its characteristics and effects. This aspect is especially important in the Chilean case, but also for many countries worldwide, where the population has been vaccinated with the virion-inactivated Sinovac vaccine.

Consistent with other studies, our results emphasize that to be worried of the side-effects decreases the acceptance of SARS-CoV-2 vaccines^24,33,34^. One of our novel findings for the existing literature is that to be worried of side-effects also decreases the acceptance to vaccinate children, while it does not affect the willingness to accept the booster dose and annual vaccination programs. We also evidence that determinants of willingness to accept vaccination vary between genders and between cohorts. For instance, trust in scientists and medical professionals influences the willingness to accept SARS-CoV-2 vaccine and booster dose only among men and among adults, but not among women or young individuals. The reasons for the gender difference in the trust in scientists and medical professionals are not clear. This can be explained by trust; however, it is not enough to explain the willingness to be vaccinated by women, which should be explored and deepened in further research. Our findings must be considered in light of some limitations, which we address here. First, we captured different degrees of confidence through an ordinal scale with 4 values, while the variability of confidence may be higher. Second, although we approached different sociodemographic characteristics, it is necessary to consider additional aspects such as cultural, ethnic, rurality and income, among others. Third, given the context of physical distance, the application of the online survey mostly reached those who have constant access to the internet and technological resources, thus excluding people with lower accessibility or knowledge about the internet and technology. Although in these limitations we highlight aspects that must be considered and remedied in future studies, it is necessary to clarify that the present study is consistent and clearly underlined the conditions that propitiate the willingness to accept an annual vaccination, booster doses and the vaccination of children. In conclusion, our study reports a high intention of the Chilean population to receive a booster dose of the COVID-19 vaccine, annual vaccination and vaccination in children under 16 years of age. Vaccine acceptance is associated with trust in scientists, medical teams and in different media such as television and radio. Perception of risk is a factor that determines in adults the acceptances of vaccination or children vaccination or booster doses.

This study provides the foundations on aspects such as trust and confidence towards COVID-19 vaccination. Other aspects such as information needs, strategies of communications and social media, or interlocutors need to be deepened in future qualitative studies, in which it might be relevant to differentiate the perception of risk in relation to COVID-19 and knowledge of it. The successful acceptance of the vaccination process against COVID-19 in the Chilean population may be useful to establish future strategies, information pathways and vaccination processes that increase the acceptance of vaccines in the population. In this framework, it is essential to continue with investigations that allow to clearly reveal the differences in trust and risk perceptions that are related to COVID-19 vaccination, addressing the subjective particularities that underlie these problems and addressing in a particular way the groups with the greatest reluctance.

## Methods

Ethical approval for this study was obtained by Universidad Autónoma de Chile ethics committee on 19 April 2021 with reference CEC 10-21. The study has a cross-sectional online survey design of adults living in Chile. A structured questionnaire was designed, validated by three experts, which was applied at the national level, in order to ensure the representativeness of the different social realities in Chile. A three-step questionnaire validation process was carried out: (a) content validity evaluation by experts (immunology, public health, design and preparation of health questionnaires); (b) construct validity, through a factor analysis of the different items included in the questionnaire; and (c) reliability, including internal consistency -assessed through Cronbach’s Alpha coefficient After this process, the questions were adjusted.

The structured questionnaire was applied through an online platform between May 21 and June 21. With a confidence level of 95% and a confidence interval of 5%, it was estimated that a minimum of 500 people older than 18 years make up a representative sample at the national level. A total of 744 volunteers completed the questionnaire, which were distributed 58% in the Metropolitan region (which concentrates 40% of the national population) and 42% in the rest of the country.

The questionnaire captured variables about willing to accept vaccines, variables of trust, variables of perceptions and sociodemographic variables. (a) Outcome variables: Willing to accept. We captured four variables related to willingness to accept vaccines: (i) Willingness to accept SARS-CoV-2 vaccine, captured in a 3-values ordinal scale (3=yes, 2=may be, and 1=no); (ii) Willingness to accept a vaccine booster shot after having a complete vaccination scheme, as a dichotomous variables (yes=1); (iii) Willingness to accept the annual vaccination and (iv) Willingness to accept the vaccination of children, last both assessed in a 4-values ordinal scale (from 1=totally disagree to 4=totally agree).

(b) explanatory variables: Trusts and perceptions.

We focused on trust (i) vaccines, (ii) stakeholders, (iii) social media, and (iv) press. To capture the trust in vaccines, the aim was to prompt people to report their trust in each of the vaccines: Sinovac, Pfizer, Cancino, AstraZeneca, Sputnik, and Johnson. We also assessed the trust in different stakeholders, such as scientists, medical professionals, policy makers, religious leaders, relatives and friends. Similarly, we included questions to assess the trust in social media as sources of information, which might influence the willingness to accept vaccines and treatments. Among social media, we asked for the trust in twitter, facebook, instagram, tiktok, whatsapp, and general websites. Last, trust in the press was also included in the questionnaire as formal information sources that might influence individuals, such as national and international newscasts, national and international newspapers, and radio broadcasts. For all questions, we used a 4-value ordinal scale in all questions about trust (from 1=no trust to 4=high trust).

In determining the association to perception variables, the aim was to prompt people to give their thinking about a set of questions about perceptions. First, how individuals perceive the effectiveness of practices to prevent COVID-19 infections was captured through a 4-value ordinal scale, from 1=no effective to 4=high effective. We included the most frequent Chilean prevention practices, such as vaccination, lockdown periods, use of masks, hand washing, social distancing, avoiding meetings, quarantine, and sanitary rooms. Second, we also captured the individual perceived risk of infection in an ordinal scale with four values (from 1=no probable to 4=highly probable). Third, the individual worry on side effects was assessed by a self-reported level of concern using a scale from no-worried (=1) to highly worried (=4). Fourth, we also assessed the perceived understanding of the vaccines of Sinovac, Pfizer, CanSino, AstraZeneca, Sputnik, Johnson, where individuals reported the level of information they considered to have about each vaccine. The individual self-report was assessed using a 4-value ordinal scale (from 1=no information to 4=high information). Fifth, the individual perception about the plausible relaxation of prevention practices due to vaccination was proxied with the claiming “the vaccination would allow” (i) to relax the use of mask, (ii) to reduce the social distancing, (iii) to avoid COVID-19 infections, (iv) to avoid the severity of COVID-19 illness, and (v) to stop the pandemic. The answer to each practice was captured in a 5-value ordinal scale (from 1=totally disagree and 5=totally agree). Sixth, we evaluate individuals’ perceived impact of the COVID-19 pandemic in their quality of life. We proxied the concept of quality of life with the impact on the job, education, health, income, familial coexistence, and general well-being of the household members. We assessed the impact of the pandemic in a 5-value ordinal scale, from 1=totally negative to 5=totally positive.

### Control variables

Last, the structured questionnaire included a set of questions to capture control variables that might be associated to the outcome or explanatory variables. We asked whether the respondent or any family member were infected with COVID-19, and whether the illness was acute. We also collected information of age, gender, administrative region of residence, schooling and nationality.

### Statistical analysis

Factor analyses followed by Cronbach’s Alpha were carried out to analyze the retained factors and the degree of internal consistency and reliability among variables of trust and perception. We took the average of each factor to create the variables of trust to be included in the multivariate regression analyses.

We were interested in estimating the association between outcome variables of willingness to accept vaccination and explanatory variables of trust and perceptions, while controlling for age, gender, and schooling. We used the following general equation:

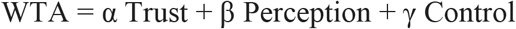

Where WTA represents the one of the four outcome variables of Willingness to Accept. Trust includes the explanatory variables of trust, while Perception stands for variables of perceptions. Last, Control captures the control variables such as (a) age, (b) gender, (c) Schooling, (d) get sick with COVID-19, (e) region of residence, and (f) nationality. α, β, and γ represent the odd ratio as a measure of association between variables.

When the outcome variables were willingness to accept SARS-CoV-2 vaccine, annual vaccines, and vaccination of children, we used ordered logistic multivariate models. In turn, logistic multivariate models were used for when the outcome variable was COVID-19 vaccine booster shot. In all models, we used the same explanatory and control variables. For each outcome variable, we adjusted a set of different models omitting one or more explanatory and control variables. We used the Akaike information criterion (AIC) to perform model comparisons and select the model with the best goodness of fit and parsimony. We ranked the AIC values and defined that the lower AIC value represents the best fitted model (Table S3).

## Data Availability

All data produced in the present study are available upon reasonable request to the authors

## Acknowledgements

This work was funded by the Vicerrectoría de Investigación y Postgrado, Universidad Autónoma de Chile grant DIUA228-202. G. Cruzat was supported by educational grant IC-UA-2021 from the Vicerrectoría de Investigación y Postgrado, Universidad Autónoma de Chile. The funders have/had no role in study design, data collection and analysis, decision to publish or preparation of the manuscript.

## Author contributions

LFF obtained ethical approval for the study via the Universidad Autónoma de Chile ethics committee. LFF, FZR, NCM and DTA conceptualized and designed the study and FZR, AA and LFF designed and performed the statistical analyses. All authors contributed to questionnaire design. CP and GC created all tables and figures for publication. LFF, NCM, DTA, AA and FZR wrote the final manuscript. All authors reviewed the results and approved the final version of the manuscript.

