## Supplemental information for "Underlying factors that influence the acceptance of COVID-19 vaccine in a country with a high vaccination rate"

**Supplementary information**

**Table S1. Summary of socio-demographic characteristics of respondents included in this study (N=744).**

| **Socio demographic characteristics** | | |
| --- | --- | --- |
| Variable | | N (%) |
| Age | 18-29 | 206 (27.7) |
|  | 30-59 | 503 (67.6) |
|  | > 59 | 35 (4.7) |
| Gender | Female | 484 (65.0) |
| Education* | Middle School | 5 (0.7) |
|  | High School | 273 (36.7) |
|  | Undergraduate | 311 (41.8) |
|  | Postgraduate | 155 (20.8) |
| Location | Capital of Chile | 433 (58.2) |
| Nationality | Chilean | 725 (97.5) |
| Infected with COVID-19 | Yes | 94 (12.6) |

*Last complete educational level

**Table S2. Milestones that occurred during the data collection period, according to the development of the pandemic and vaccination in Chile.**

| **DATE** | **EVENT** |
| --- | --- |
| **May 21** | The highest incidence rate in Chile reaches 12.7% |
| **May 25** | Vaccination begins in people younger than 30 years |
| **May 27** | 6% increase in new cases nationwide compared to the last seven days |
| **May 31** | 2020 Public Account of the Chilean Ministry of Health. |
| **June 2** | The highest incidence rate in Chile reaches 11.9% |
| **June 3** | Mobility Pass permits are restricted |
| **June 4** | Chile achieved 15,053,577 PCR examinations, which places our country in a leadership position in Latin America and the world. The positivity in the last 24 hours is 10%. |
| **June 7** | Chilean Ministry of Health reports that 75% of new COVID-19 cases haven’t completed their vaccination schedule |
| **June 10** | All the communes of the capital of Chile go into lockdown. |
| **June 11** | Chilean Ministry of Health announces the imminent start of vaccination in children between 17 and 12 years old with Pfizer vaccine |
| **June 13** | Elections of Regional Governors are held |
| **June 15** | 11 regions decreased their cases in the last seven days and 10 in the last 14 day. |
| **June 17** | 11% decrease in cases in one week. |
| **June 19** | Conversation table between Chilean Ministry of Health and different guild and specialists to review health strategies. |
| **June 20** | Strategy proposed by the Medical College on "Epidemiological circuit-breaker" is rejected |
| **June 21** | 24 of 52 communes of capital of Chile are released from lockdown |

**Table S3. Ranking of models to estimate the association of the outcome variables of willingness to accept (a) SARS-CoV-2 vaccine, (b) third dose, (c) annual vaccination, and (d) vaccine to children, against explanatory variables of trust and perceptions. Models varied in the number of variables of trust and perceptions. k is the number of parameters in the model, including outcome and explanatory variables. AICc refers to Akaike’s Information Criterion adjusted for small sample size and ⊗ AICc is the difference in AICc value of each model from the core model. Model were grouped according to AICc value in Core model (smallest value), Candidate models (⊗ AICc <2), and Models with relatively little support (⊗ AICc>2). Explanatory variables: [1] Trust in vaccines, [2] Trust in scientists and medical professionals, [3] Trust in policy makers, [4] Trust in religious leaders, [5] Trust in relatives, [6] Trust in social media, [7] Trust in press, [8] Perceived effectiveness of prevention practices, [9] Perceived risk infection, [10] Worry on side effects, [11] Perceived understanding on vaccines, [12] Perceived prevention of severity by vaccines, [13] Perceived relaxation of prevention practices thanks to vaccination, [14] Perceived pandemic stopping thanks to vaccination, [15] Perceived impact in quality of life, [16] Infection by covid-19 in family, [17] Age, [18] Gender, [19] Schooling, [20] get sick with COVID-19, [21] region of residence, [22] acute COVID-19 infection of a family member, [23] Nationality.**

|  | Willingness to accept: | | | |  |  | | | |  |  | | | |  |  | | | |
| --- | --- | --- | --- | --- | --- | --- | --- | --- | --- | --- | --- | --- | --- | --- | --- | --- | --- | --- | --- |
|  | SARS-CoV-2 vaccine | | | |  | third dose | | | |  | annual vaccination | | | |  | vaccine to children | | | |
| Models | Omitted variables | k | AICc | ⊗ AICc |  | Omitted variables | k | AICc | ⊗ AICc |  | Omitted variables | k | AICc | ⊗ AICc |  | Omitted variables | k | AICc | ⊗ AICc |
| Core model | [20] to [23] | 19 | 281.5 | 0.0 |  | [20] to [23] | 19 | 348.2 | 0.0 |  | [20] to [23] | 19 | 1287.3 | 0.0 |  | [20] to [23] | 21 | 1096.7 | 0.0 |
| Candidate models | [20] to [22] | 20 | 283.4 | 1.9 |  | [20] to [22] | 20 | 349.7 | 1.4 |  | [20] to [22] | 20 | 1288.3 | 1.0 |  | [21], [23] | 19 | 1096.8 | 0.1 |
|  |  |  |  |  |  |  |  |  |  |  | [20], [23] | 21 | 1288.9 | 1.6 |  | [23] | 22 | 1097.5 | 0.8 |
|  |  |  |  |  |  |  |  |  |  |  | [16], [20] | 21 | 1288.9 | 1.6 |  | [22] | 22 | 1097.9 | 1.3 |
|  |  |  |  |  |  |  |  |  |  |  | [20], [21] | 21 | 1288.9 | 1.6 |  | [16] | 22 | 1097.9 | 1.3 |
|  |  |  |  |  |  |  |  |  |  |  | [20], [22] | 21 | 1289.0 | 1.7 |  | [21] | 22 | 1098.5 | 1.8 |
|  |  |  |  |  |  |  |  |  |  |  | [21], [23] | 21 | 1289.0 | 1.7 |  | [20], [23] | 21 | 1098.6 | 1.9 |
|  |  |  |  |  |  |  |  |  |  |  | [16] | 22 | 1289.2 | 1.8 |  |  |  |  |  |
| Models with | [20], [22] | 21 | 285.5 | 4.0 |  | [20], [21] | 21 | 351.6 | 3.4 |  | [22] | 22 | 1289.7 | 2.4 |  | [20] to [22] | 20 | 1098.8 | 2.1 |
| relatively | [20], [21] | 21 | 285.5 | 4.0 |  | [16], [20] | 21 | 351.6 | 3.4 |  | [23] | 22 | 1289.7 | 2.4 |  | [20], [22] | 21 | 1099.3 | 2.6 |
| little support | [16], [20] | 21 | 285.5 | 4.0 |  | [20], [22] | 21 | 351.8 | 3.5 |  | [20] | 22 | 1289.7 | 2.4 |  | - | 23 | 1099.4 | 2.7 |
|  | [21], [23] | 21 | 285.6 | 4.1 |  | [21], [23] | 21 | 352.2 | 3.9 |  | [21] | 22 | 1289.9 | 2.6 |  | [16], [20] | 21 | 1099.9 | 3.3 |
|  | [20], [23] | 21 | 285.7 | 4.2 |  | [20], [23] | 21 | 352.3 | 4.0 |  | - | 23 | 1290.4 | 3.1 |  | [20], [21] | 21 | 1100.1 | 3.5 |
|  | [22] | 22 | 287.5 | 6.0 |  | [21] | 22 | 353.6 | 5.4 |  |  |  |  |  |  | [20] | 22 | 1100.5 | 3.9 |
|  | [21] | 22 | 287.5 | 6.0 |  | [20] | 22 | 353.7 | 5.5 |  |  |  |  |  |  |  |  |  |  |
|  | [16] | 22 | 287.6 | 6.1 |  | [16] | 22 | 353.7 | 5.5 |  |  |  |  |  |  |  |  |  |  |
|  | [20] | 22 | 287.6 | 6.1 |  | [22] | 22 | 353.8 | 5.6 |  |  |  |  |  |  |  |  |  |  |
|  | [23] | 22 | 287.7 | 6.2 |  | [23] | 22 | 354.3 | 6.1 |  |  |  |  |  |  |  |  |  |  |
|  | - | 23 | 289.6 | 8.1 |  | - | 23 | 355.8 | 7.5 |  |  |  |  |  |  |  |  |  |  |

**Table S4. Associations of the willingness to SARS-CoV-2 vaccination, third dose, annual vaccination, and to vaccinate children, with variables of trust and perception among Chilean female (n=484) and male individuals (n=260).**

|  |  | Willingness to SARS-CoV-2 vaccination | |  | Willingness to third dose vaccination | |  | Willingness to annual vaccination | |  | Willingness to vaccinate children | |
| --- | --- | --- | --- | --- | --- | --- | --- | --- | --- | --- | --- | --- |
|  |  | Female | Men |  | Female | Men |  | Female | Men |  | Female | Men |
| Explanatory variables |  | [1] | [2] |  | [3] | [4] |  | [5] | [6] |  | [7] | [8] |
| Trust in vaccines | [a] | 4.8** | 5.8 |  | 4.0** | 3.4 |  | 2.4** | 1.8* |  | 2.1** | 1.8 |
|  |  | (2.1 - 11.0) | (0.7 - 50.0) |  | (1.9 - 8.2) | (0.9 - 12.1) |  | (1.6 - 3.5) | (1.1 - 3.2) |  | (1.4 - 3.2) | (1.0 - 3.2) |
| Trust in scientists and medical professionals | [b] | 2.2 | 46.1* |  | 1.9 | 4.2* |  | 2.0** | 2.5** |  | 2.0** | 3.8** |
|  |  | (0.9 - 5.2) | (2.5 - 862.1) |  | (0.9 - 3.9) | (1.3 - 13.1) |  | (1.3 - 3.0) | (1.4 - 4.4) |  | (1.3 - 3.0) | (2.1 - 6.9) |
| Trust in policy makers | [c] | 3.1* | 0.6 |  | 2.4* | 1.0 |  | 1.2 | 1.2 |  | 1.3 | 1.1 |
|  |  | (1.2 - 8.2) | (0.0 - 18.3) |  | (1.1 - 5.3) | (0.3 - 2.9) |  | (0.9 - 1.7) | (0.7 - 2.0) |  | (0.9 - 1.9) | (0.6 - 2.1) |
| Trust in religious leaders | [d] | 0.9 | 2.0 |  | 0.8 | 0.4 |  | 0.8 | 0.7 |  | 0.7* | 0.6* |
|  |  | (0.4 - 2.1) | (0.0 - 111.8) |  | (0.4 - 1.6) | (0.2 - 1.0) |  | (0.6 - 1.1) | (0.5 - 1.1) |  | (0.5 - 1.0) | (0.4 - 0.9) |
| Trust in relatives | [e] | 1.2 | 8.0 |  | 1.7 | 1.3 |  | 1.1 | 1.1 |  | 1.0 | 1.8* |
|  |  | (0.6 - 2.8) | (0.6 - 112.6) |  | (0.8 - 3.7) | (0.5 - 3.3) |  | (0.8 - 1.5) | (0.7 - 1.7) |  | (0.7 - 1.4) | (1.1 - 2.9) |
| Trust in social media | [f] | 1.1 | 0.1 |  | 0.3** | 0.5 |  | 1.2 | 0.8 |  | 0.7 | 0.7 |
|  |  | (0.4 - 2.7) | (0.0 - 1.3) |  | (0.1 - 0.7) | (0.2 - 1.3) |  | (0.8 - 1.8) | (0.5 - 1.3) |  | (0.4 - 1.0) | (0.4 - 1.2) |
| Trust in press | [g] | 0.5 | 12.0 |  | 2.3 | 1.1 |  | 1.1 | 1.1 |  | 1.3 | 1.0 |
|  |  | (0.2 - 1.4) | (0.4 - 343.8) |  | (1.0 - 5.6) | (0.4 - 3.3) |  | (0.7 - 1.6) | (0.7 - 1.8) |  | (0.9 - 2.0) | (0.6 - 1.8) |
| Perceived effectiveness of prevention practices | [h] | 3.2* | 11.1 |  | 4.3** | 1.8 |  | 3.1** | 1.8* |  | 3.9** | 1.4 |
|  |  | (1.1 - 9.5) | (0.9 - 133.3) |  | (1.7 - 11.3) | (0.6 - 5.1) |  | (1.8 - 5.1) | (1.0 - 3.2) |  | (2.3 - 6.7) | (0.7 - 2.5) |
| Perceived risk infection | [i] | 2.4* | 1.9 |  | 1.7 | 1.1 |  | 1.3 | 1.4 |  | 1.2 | 1.0 |
|  |  | (1.1 - 5.1) | (0.3 - 13.4) |  | (0.9 - 3.1) | (0.5 - 2.5) |  | (0.9 - 1.8) | (0.9 - 2.2) |  | (0.9 - 1.7) | (0.6 - 1.6) |
| Worry on side effects | [j] | 0.5** | 0.2* |  | 1.1 | 0.6 |  | 1.0 | 0.8 |  | 0.9 | 0.7* |
|  |  | (0.3 - 0.8) | (0.0 - 0.9) |  | (0.7 - 1.8) | (0.3 - 1.2) |  | (0.8 - 1.2) | (0.6 - 1.1) |  | (0.7 - 1.1) | (0.5 - 0.9) |
| Perceived understanding on vaccines | [k] | 0.8 | 1.1 |  | 0.9 | 0.8 |  | 1.1 | 1.2 |  | 1.0 | 1.3 |
|  |  | (0.4 - 1.7) | (0.1 - 8.5) |  | (0.4 - 1.9) | (0.3 - 2.4) |  | (0.8 - 1.7) | (0.7 - 1.9) |  | (0.7 - 1.5) | (0.8 - 2.3) |
| Perceived prevention of severity by vaccines | [l] | 1.0 | 35.0** |  | 1.0 | 1.0 |  | 1.0 | 1.0 |  | 0.8* | 0.7* |
|  |  | (0.6 - 1.5) | (2.7 - 462.4) |  | (0.7 - 1.5) | (0.6 - 1.7) |  | (0.8 - 1.2) | (0.8 - 1.3) |  | (0.7 - 1.0) | (0.6 - 1.0) |
| Perceived relaxation of prevention practices thanks to vaccination | [m] | 1.3 | 3.5 |  | 0.9 | 0.6 |  | 0.7** | 0.7 |  | 1.2 | 0.9 |
|  |  | (0.7 - 2.3) | (0.5 - 23.7) |  | (0.5 - 1.5) | (0.3 - 1.2) |  | (0.5 - 0.9) | (0.5 - 1.0) |  | (0.9 - 1.6) | (0.6 - 1.3) |
| Perceived pandemic stopping thanks to vaccination | [n] | 1.0 | 14.6** |  | 1.2 | 2.1* |  | 1.4** | 1.6** |  | 1.2* | 1.5** |
|  |  | (0.6 - 1.6) | (1.9 - 111.7) |  | (0.8 - 1.8) | (1.2 - 3.9) |  | (1.2 - 1.7) | (1.2 - 2.1) |  | (1.0 - 1.5) | (1.1 - 2.0) |
| Perceived impact in quality of life | [o] | 0.7 | 0.1 |  | 1.3 | 0.5 |  | 1.0 | 0.6* |  | 0.9 | 0.7 |
|  |  | (0.3 - 1.3) | (0.0 - 1.5) |  | (0.7 - 2.4) | (0.2 - 1.1) |  | (0.7 - 1.4) | (0.4 - 1.0) |  | (0.6 - 1.3) | (0.4 - 1.0) |
| Infection by covid-19 in family | [p] | 0.4 | 31.7 |  | 0.7 | 2.2 |  | 1.1 | 1.0 |  | 0.7 | 1.0 |
|  |  | (0.2 - 1.1) | (0.4 - 2,465.2) |  | (0.3 - 1.5) | (0.5 - 8.5) |  | (0.7 - 1.7) | (0.6 - 1.9) |  | (0.5 - 1.2) | (0.5 - 2.0) |
| Age | [q] | 1.1* | 1.2* |  | 1.0 | 1.0 |  | 1.0 | 1.0 |  | 1.0** | 1.0** |
|  |  | (1.0 - 1.1) | (1.0 - 1.4) |  | (0.9 - 1.0) | (1.0 - 1.1) |  | (1.0 - 1.0) | (1.0 - 1.0) |  | (1.0 - 1.1) | (1.0 - 1.1) |
| Schooling | [r] | 1.0 | 0.5 |  | 0.9 | 1.0 |  | 0.9 | 1.0 |  | 0.9 | 0.8 |
|  |  | (0.7 - 1.4) | (0.2 - 1.1) |  | (0.7 - 1.2) | (0.7 - 1.5) |  | (0.8 - 1.0) | (0.8 - 1.2) |  | (0.8 - 1.1) | (0.7 - 1.0) |
| Multivariate model |  | Ordered logit | Ordered logit |  | Logit | Logit |  | Ordered logit | Ordered logit |  | Ordered logit | Ordered logit |
| Observations |  | 484 | 260 |  | 484 | 260 |  | 484 | 260 |  | 484 | 260 |

Note= Columns [1], [2], [5], [6], [7], and [8] show results of ordered logit multivariate models. Column [3] and [4] show logit model results. For all columns, cells show odd ratio coefficients and, in parenthesis, confidence intervals at 95%. For each outcome variable, table shows the model with best goodness of fit and parsimony compared with other candidate models, which was selected using Akaike Information Criterion (see Table S3). * and ** refer to significant levels at 5% and 1%.

**Table S5. Associations of the willingness to SARS-CoV-2 vaccination, third dose, annual vaccination, and to vaccinate children, with variables of trust and perception among Chilean young (<30 years old, n=206) and adult individuals (>29 and <60 years old, n=503).**

|  |  | Willingness to SARS-CoV-2 vaccination | |  | Willingness to third dose vaccination | |  | Willingness to annual vaccination | |  | Willingness to vaccinate children | |
| --- | --- | --- | --- | --- | --- | --- | --- | --- | --- | --- | --- | --- |
|  |  | Young | Adult |  | Young | Adult |  | Young | Adult |  | Young | Adult |
| Explanatory variables |  | [1] | [2] |  | [3] | [4] |  | [5] | [6] |  | [7] | [8] |
| Trust in vaccines | [a] | 6.0** | 3.4* |  | 9.9** | 2.2* |  | 2.3** | 1.9** |  | 2.8** | 1.5 |
|  |  | (1.6 - 21.7) | (1.1 - 10.0) |  | (2.4 - 41.6) | (1.0 - 4.9) |  | (1.4 - 3.9) | (1.3 - 2.9) |  | (1.6 - 4.8) | (1.0 - 2.4) |
| Trust in scientists and medical professionals | [b] | 2.9 | 3.7* |  | 1.2 | 3.9** |  | 1.6 | 2.6** |  | 1.1 | 3.9** |
|  |  | (0.7 - 12.6) | (1.2 - 11.8) |  | (0.3 - 4.4) | (1.8 - 8.5) |  | (0.8 - 3.0) | (1.7 - 3.9) |  | (0.6 - 2.2) | (2.5 - 6.2) |
| Trust in policy makers | [c] | 3.3 | 4.9* |  | 1.5 | 2.1 |  | 1.2 | 1.2 |  | 2.1* | 1.2 |
|  |  | (0.7 - 15.3) | (1.3 - 18.5) |  | (0.4 - 5.4) | (1.0 - 4.5) |  | (0.7 - 2.1) | (0.8 - 1.7) |  | (1.2 - 3.8) | (0.8 - 1.8) |
| Trust in religious leaders | [d] | 1.8 | 0.7 |  | 0.5 | 0.5* |  | 0.6* | 0.8 |  | 0.4** | 0.7 |
|  |  | (0.3 - 9.3) | (0.3 - 1.7) |  | (0.1 - 1.6) | (0.3 - 1.0) |  | (0.3 - 1.0) | (0.6 - 1.1) |  | (0.2 - 0.8) | (0.5 - 1.0) |
| Trust in relatives | [e] | 0.4 | 3.9** |  | 6.0* | 1.2 |  | 1.1 | 1.1 |  | 1.1 | 1.3 |
|  |  | (0.1 - 1.4) | (1.5 - 9.9) |  | (1.3 - 28.7) | (0.6 - 2.3) |  | (0.6 - 2.0) | (0.8 - 1.6) |  | (0.6 - 2.0) | (0.9 - 1.9) |
| Trust in social media | [f] | 3.6 | 0.6 |  | 0.2* | 0.3** |  | 1.1 | 0.9 |  | 0.7 | 0.7 |
|  |  | (0.7 - 17.0) | (0.2 - 1.8) |  | (0.1 - 1.0) | (0.2 - 0.7) |  | (0.6 - 2.1) | (0.7 - 1.3) |  | (0.4 - 1.3) | (0.5 - 1.0) |
| Trust in press | [g] | 0.4 | 0.7 |  | 1.9 | 1.3 |  | 1.0 | 1.1 |  | 1.4 | 1.0 |
|  |  | (0.1 - 2.1) | (0.2 - 2.4) |  | (0.5 - 7.6) | (0.6 - 2.9) |  | (0.6 - 1.8) | (0.8 - 1.6) |  | (0.8 - 2.6) | (0.6 - 1.5) |
| Perceived effectiveness of prevention practices | [h] | 2.9 | 2.3 |  | 6.5 | 2.1 |  | 4.7** | 1.9** |  | 2.8* | 2.3** |
|  |  | (0.4 - 21.8) | (0.9 - 5.9) |  | (0.8 - 53.5) | (1.0 - 4.2) |  | (1.9 - 11.3) | (1.2 - 2.9) |  | (1.2 - 6.9) | (1.4 - 3.6) |
| Perceived risk infection | [i] | 3.6 | 1.3 |  | 1.7 | 1.3 |  | 1.3 | 1.3 |  | 1.3 | 1.1 |
|  |  | (1.0 - 13.2) | (0.6 - 3.1) |  | (0.5 - 5.2) | (0.8 - 2.3) |  | (0.8 - 2.2) | (1.0 - 1.8) |  | (0.8 - 2.2) | (0.8 - 1.6) |
| Worry on side effects | [j] | 0.3** | 0.7 |  | 0.3* | 0.9 |  | 0.9 | 0.9 |  | 0.8 | 0.7* |
|  |  | (0.1 - 0.6) | (0.3 - 1.3) |  | (0.1 - 0.8) | (0.6 - 1.5) |  | (0.6 - 1.3) | (0.7 - 1.1) |  | (0.6 - 1.2) | (0.6 - 0.9) |
| Perceived understanding on vaccines | [k] | 1.5 | 0.4* |  | 0.7 | 0.6 |  | 2.1* | 0.9 |  | 1.6 | 0.9 |
|  |  | (0.3 - 6.5) | (0.2 - 1.0) |  | (0.2 - 2.3) | (0.3 - 1.2) |  | (1.1 - 3.8) | (0.6 - 1.2) |  | (0.9 - 3.0) | (0.6 - 1.4) |
| Perceived prevention of severity by vaccines | [l] | 0.8 | 2.3** |  | 1.8 | 0.9 |  | 0.9 | 1.0 |  | 0.7* | 0.9 |
|  |  | (0.4 - 1.6) | (1.3 - 4.1) |  | (0.9 - 3.9) | (0.6 - 1.2) |  | (0.7 - 1.3) | (0.8 - 1.2) |  | (0.5 - 1.0) | (0.7 - 1.0) |
| Perceived relaxation of prevention practices thanks to vaccination | [m] | 1.1 | 1.7 |  | 0.4 | 0.9 |  | 0.8 | 0.7** |  | 0.7 | 1.2 |
|  |  | (0.4 - 3.0) | (0.9 - 3.4) |  | (0.1 - 1.3) | (0.6 - 1.4) |  | (0.5 - 1.2) | (0.6 - 0.9) |  | (0.4 - 1.1) | (0.9 - 1.5) |
| Perceived pandemic stopping thanks to vaccination | [n] | 0.9 | 2.0* |  | 2.4 | 1.3 |  | 1.4* | 1.5** |  | 1.3 | 1.4** |
|  |  | (0.4 - 2.1) | (1.1 - 3.6) |  | (1.0 - 5.6) | (0.9 - 1.8) |  | (1.0 - 1.8) | (1.3 - 1.8) |  | (0.9 - 1.7) | (1.2 - 1.8) |
| Perceived impact in quality of life | [o] | 0.6 | 0.4* |  | 0.1* | 1.1 |  | 0.8 | 0.8 |  | 0.7 | 0.8 |
|  |  | (0.2 - 2.1) | (0.2 - 0.8) |  | (0.0 - 0.7) | (0.6 - 1.9) |  | (0.5 - 1.4) | (0.6 - 1.1) |  | (0.4 - 1.3) | (0.6 - 1.1) |
| Infection by covid-19 in family | [p] | 0.2 | 0.7 |  | 0.2 | 0.9 |  | 1.2 | 0.9 |  | 0.4* | 0.9 |
|  |  | (0.0 - 1.2) | (0.2 - 2.4) |  | (0.0 - 1.2) | (0.4 - 1.9) |  | (0.6 - 2.5) | (0.6 - 1.4) |  | (0.2 - 0.9) | (0.5 - 1.3) |
| Age | [q] | 1.5* | 1.0 |  | 1.0 | 1.0 |  | 0.9 | 1.0 |  | 1.1 | 1.1** |
|  |  | (1.1 - 2.2) | (0.9 - 1.1) |  | (0.8 - 1.4) | (0.9 - 1.0) |  | (0.8 - 1.1) | (1.0 - 1.0) |  | (1.0 - 1.2) | (1.0 - 1.1) |
| Gender (female=1) | [r] | 1.8 | 1.6 |  | 3.6 | 1.2 |  | 1.6 | 1.1 |  | 1.3 | 1.5 |
|  |  | (0.2 - 13.3) | (0.5 - 5.4) |  | (0.5 - 26.1) | (0.5 - 2.6) |  | (0.7 - 3.4) | (0.7 - 1.7) |  | (0.6 - 3.0) | (1.0 - 2.5) |
| Schooling | [s] | 0.8 | 0.9 |  | 0.9 | 1.0 |  | 1.0 | 0.9 |  | 1.0 | 0.8** |
|  |  | (0.5 - 1.4) | (0.6 - 1.3) |  | (0.5 - 1.4) | (0.8 - 1.3) |  | (0.8 - 1.3) | (0.8 - 1.1) |  | (0.8 - 1.3) | (0.7 - 0.9) |
| Multivariate model |  | Ordered logit | Ordered logit |  | Logit | Logit |  | Ordered logit | Ordered logit |  | Ordered logit | Ordered logit |
| Observations |  | 206 | 503 |  | 206 | 503 |  | 206 | 503 |  | 206 | 503 |

Notes

Note 1= Elderly persons (>59 years old) were omitted from models because the sample (n=35).

Note 2= Columns [1], [2], [5], [6], [7], and [8] show results of ordered logit multivariate models. Column [3] and [4] show logit model results. For all columns, cells show odd ratio coefficients and, in parenthesis, confidence intervals at 95%. For each outcome variable, table shows the model with best goodness of fit and parsimony compared with other candidate models, which was selected using Akaike Information Criterion (see Table S3). * and ** refer to significant levels at 5% and 1%.

**Questionnaire**

**VACCINE ACCEPTANCE SURVEY**

**“VACCINE AGAINST SARS-COV-2 IN CHILE: UNDERSTANDING THE DETERMINANTS ASSOCIATED WITH THE POPULATION’S WILLINGNESS TO ACCEPT IT”**

A1. What age are you? ___________
A2. What is your gender?
1. Male
2. Female
3. Other
A3. What is your nationality?
1. Chilean
2. Peruvian
3. Colombian
4. Venezuelan
5. Bolivian
6. Haitian
7. Argentinian
8. Dominican
9. Other ¿Which? ___ **OPEN TEXT BOX ANCHORED, MAX 250 CHARACTERS**

Does not know – Does not answer

A4. In what Region do you live?
1. I Region (Tarapacá)
2. II Region (Antofagasta)
3. III Region (Atacama)
4. IV Region (Coquimbo)
5. V Region (Valparaíso)
6. VI Region (Libertador Bernardo O’Higgins)
7. VII Region (Maule)
8. VIII Region (Biobío)
9. IX Region (La Araucanía)
10. X Region (Los Lagos)
11. XI Region (Aysén)
12. XII Region (Magallanes)
13. Metropolitan Region
14. XIV Region (Los Ríos)
15. XV Region (Arica y Parinacota)
16. XVI Region (Ñuble)
A5. ZONE

| North | Center | South | RM |
| --- | --- | --- | --- |
| 1 | 2 | 3 | 4 |

A6. **PLEASE INSERT NSE CHILE**
A7. What is your comunne of residence?
A8. What is your last educational level?
1. Incomplete primary school
2. Complete primary school
3. Incomplete secondary school
4. Complete secondary school
5. Incomplete technical studies
6. Complete technical studies
7. Incomplete undergraduate studies
8. Complete undergraduate studies
9. Postgraduate studies
A9. Do you have children under 16 years old?

1. Yes

2. No

A10. What choice better represents your current situation about the vaccine?
1. Already vaccinated 1st or 2nd dose
2. Waiting to get vaccinated
3. Does not decide if will get vaccinated
4. Will not get vaccinated

**QUESTIONS ABOUT COVID-19 SEVERITY**

A11. Have you gotten sick of COVID-19?
1. No
2. Yes, asymptomatic
3. Yes, symptomatic, no hospitalization
4. Yes, hospitalized
A12. Has a member of your family (father, mother, siblings, kids) gotten sick of COVID-19?
1. No **-**> **SKIP TO A16**
2. Yes, asymptomatic
3. Yes, symptomatic, no hospitalization
4. Yes, hospitalized
5. Yes, died
A13. Did the person that got COVID-19 live with you?
1. Yes
2. No
A14. How likely do you think it is to contract and get sick with COVID-19?
1. Unlikely
2. Less likely
3. Likely
4. Highly likely

**TRUST IN THE SOURCE OF VACCINE INFORMATION**

A15. How much do you trust in COVID-19 vaccine information provided by?

|  |  | No trust | Little trust | Trust | High trust |
| --- | --- | --- | --- | --- | --- |
| 1 | Family | 1 | 2 | 3 | 4 |
| 2 | Friends | 1 | 2 | 3 | 4 |
| 3 | Religious leaders | 1 | 2 | 3 | 4 |
| 4 | Politicians | 1 | 2 | 3 | 4 |
| 5 | Ministry of Health | 1 | 2 | 3 | 4 |
| 6 | Medical college professionals | 1 | 2 | 3 | 4 |
| 7 | Scientists | 1 | 2 | 3 | 4 |
| 8 | ISP (Instituto de Salud Pública) profesional | 1 | 2 | 3 | 4 |
| 9 | WHO (World Health Organization) professionals | 1 | 2 | 3 | 4 |

A16. How much do you trust the information about COVID-19 vaccine provided by?

|  |  | No trust | Little trust | Trust | High trust | Do not use that media |
| --- | --- | --- | --- | --- | --- | --- |
| 1 | Facebook | 1 | 2 | 3 | 4 | 5 |
| 2 | Instagram | 1 | 2 | 3 | 4 | 5 |
| 3 | Tik Tok | 1 | 2 | 3 | 4 | 5 |
| 4 | Twitter | 1 | 2 | 3 | 4 | 5 |
| 5 | WhatsApp | 1 | 2 | 3 | 4 | 5 |
| 6 | Websites | 1 | 2 | 3 | 4 | 5 |
| 7 | National television | 1 | 2 | 3 | 4 | 5 |
| 8 | International television | 1 | 2 | 3 | 4 | 5 |
| 9 | National newspaper | 1 | 2 | 3 | 4 | 5 |
| 10 | International newspaper | 1 | 2 | 3 | 4 | 5 |
| 11 | Radio | 1 | 2 | 3 | 4 | 5 |

**TRUST IN THE VACCINE**

A17. How worried do you feel about the possible side effects of COVID-19 vaccines?
1. Not worried
2. Little worried
3. Worried
4. Very worried
A18. When deciding whether to vaccinate or not against COVID-19, how relevant is it for you the time it took to develop a COVID-19 vaccine?

1. Not relevant

2. Little relevant

3. Relevant

4. Highly relevant

A19. According to what you know or have heard, how effective do you think the following measures are to prevent COVID-19?

|  |  | Not effective | Little effective | Effective | Highly effective | Do not know |
| --- | --- | --- | --- | --- | --- | --- |
| 1 | Vaccination | 1 | 2 | 3 | 4 | 99 |
| 2 | Quarantine | 1 | 2 | 3 | 4 | 99 |
| 3 | Use of mask | 1 | 2 | 3 | 4 | 99 |
| 4 | Washing hands | 1 | 2 | 3 | 4 | 99 |
| 5 | Keeping physical distance | 1 | 2 | 3 | 4 | 99 |
| 6 | Avoiding social meetings | 1 | 2 | 3 | 4 | 99 |
| 7 | Home isolation | 1 | 2 | 3 | 4 | 99 |
| 8 | Use of sanitary residences | 1 | 2 | 3 | 4 | 99 |

A20. How much information do you have about the following vaccines?

|  |  | No information | Little information | Enough information | A lot of information | Do not know |
| --- | --- | --- | --- | --- | --- | --- |
| 1 | Sinovac (China) | 1 | 2 | 3 | 4 | 99 |
| 2 | Pfizer (Germany/USA) | 1 | 2 | 3 | 4 | 99 |
| 3 | CanSino (China /Canada) | 1 | 2 | 3 | 4 | 99 |
| 4 | AstraZeneca (UK/Sweden) | 1 | 2 | 3 | 4 | 99 |
| 5 | Sputnik V (Russia) | 1 | 2 | 3 | 4 | 99 |
| 6 | Johnson & Johnson (USA/Belgium) | 1 | 2 | 3 | 4 | 99 |

A21. How much do you trust in the following vaccines?

|  |  | No trust | Little trust | Trust | High trust | Do not know |
| --- | --- | --- | --- | --- | --- | --- |
| 1 | Sinovac (China) | 1 | 2 | 3 | 4 | 99 |
| 2 | Pfizer (Germany/USA) | 1 | 2 | 3 | 4 | 99 |
| 3 | CanSino (China /Canada) | 1 | 2 | 3 | 4 | 99 |
| 4 | AstraZeneca (UK/Sweden) | 1 | 2 | 3 | 4 | 99 |
| 5 | Sputnik V (Russia) | 1 | 2 | 3 | 4 | 99 |
| 6 | Johnson & Johnson (USA/Bélgica) | 1 | 2 | 3 | 4 | 99 |

A22. If you would have to pay for a COVID-19 vaccine, would you get vaccinated?
1. Yes
2. No
99. Do not know
A23. In the hypothetical situation that you already have two doses of the COVID-19 vaccine and you are informed that a third dose is necessary, would you get the third dose?
1. Yes
2. No
99. Do not know
A24. If it was necessary to get vaccinated against COVID-19 every year, as is the case with the Influenza vaccine, how willing are you to get vaccinated every year?
1. Not willing to
2. Little willing to
3. Willing to
4. Highly willing to

**POST VACCINATION IMPACT**

A25. Do you agree or disagree with the following sentences?

|  |  | Highly disagree | Disagree | Do not agree or disagree | Agree | Highly agree | Do not know |
| --- | --- | --- | --- | --- | --- | --- | --- |
| 1 | With vaccination you can stop using mask | 1 | 2 | 3 | 4 | 5 | 99 |
| 2 | With vaccination the measures to prevent physical interactions with other people can be relaxed | 1 | 2 | 3 | 4 | 5 | 99 |
| 3 | The vaccine will avoid me of contract COVID-19 | 1 | 2 | 3 | 4 | 5 | 99 |
| 4 | The vaccine will only avoid me to get severe disease | 1 | 2 | 3 | 4 | 5 | 99 |
| 5 | Vaccination will stop the COVID-19 pandemic | 1 | 2 | 3 | 4 | 5 | 99 |

A26. **FILTER: (A11=1)** If a COVID-19 vaccine is approved for children under 16 years old, would you vaccinate your children?

**FILTER: (A11=2)** (If you had children under 16 years old and a COVID-19 vaccine would be approved), would you vaccinate your children?

1. Definitively no
2. Probably no
3. Maybe yes
4. Definitely yes
A27. In your experience, how much has your life been impacted because of the pandemic in the following aspects?

|  |  | Very negatively | Negatively | Not negatively not positively | Positively | Very positively |
| --- | --- | --- | --- | --- | --- | --- |
| 1 | Your wellness | 1 | 2 | 3 | 4 | 5 |
| 2 | The jobs of your household members | 1 | 2 | 3 | 4 | 5 |
| 3 | The coexistence with your household members | 1 | 2 | 3 | 4 | 5 |
| 4 | The education of your household members | 1 | 2 | 3 | 4 | 5 |
| 5 | The health of your household members | 1 | 2 | 3 | 4 | 5 |
| 6 | The income of your household members | 1 | 2 | 3 | 4 | 5 |

**THANK YOU AND FINISH**
